# Prevalence of erectile dysfunction patients with diabetes mellitus and its association with body mass index and glycated hemoglobin in Africa: a systematic review and meta-analysis

**DOI:** 10.1101/19011866

**Authors:** Wondimeneh Shibabaw Shiferaw, Tadesse Yirga Akalu, Yared Asmare Aynalem

**Affiliations:** Lecturer of Nursing, Department of Nursing, College of Health Science, Debre Berhan University Address: P.O. Box 445, Debre Berhan, Ethiopia; Lecturer of Nursing, Department of Nursing, College of Health Science, Debre markos University Address: P.O. Box 269, Debre Markos, Ethiopia

**Keywords:** erectile dysfunction, impotence, sexual dysfunction, diabetes mellitus, Africa

## Abstract

**Background:** Mortality and morbidity in patients with diabetes mellitus (DM) is attributed to both the micro-vascular and macro-vascular complications. Variation among primary studies was seen on the prevalence of erectile dysfunction in Africa. Therefore, this study was aimed to estimate the pooled prevalence of erectile dysfunction patients with diabetes mellitus and its association with body mass index and glycated hemoglobin in Africa.

**Methods:** PubMed, Web of Science, Cochrane library, Scopus, Psyinfo, Africa online journal and Google Scholar were searched. A funnel plot and Egger’s regression test were used to see publication bias. I-squared statistic was used to check heterogeneity of studies. DerSimonian and Laird random-effects model was applied to estimate the pooled effect size. The subgroup and Meta regression analysis were conducted by country, sample size, and year of publication. Sensitivity analysis was deployed to see the effect of single study on the overall estimation. STATA version 14 statistical software was used for meta-analysis.

**Result:** A total of 20 studies with 5,177 study participants were included to estimate the pooled prevalence. The pooled prevalence of erectile dysfunction patients with diabetes mellitus was 61.62% (95% CI: 48.35–74.9). BMI ≥ 30kg/m^2^ (AOR = 1.26; 95% CI: 0.73 –2.16), and glycated hemoglobin ≥ 7% (AOR = 0.93; 95% CI: 0.5–5.9), were identified factors though not statistically significant associated with erectile dysfunction.

**Conclusions:** The prevalence of erectile dysfunction in Africa remains high. Therefore, situation based interventions and country context specific preventive strategies could be developed to reduce the magnitude of erectile dysfunction among patients with diabetes mellitus.

## Introduction

Diabetes is a major public health problem, increasingly affecting millions of people across the globe [1]. About 425 million peoples are suffering from diabetes mellitus (DM) globally in 2017; by 2045 this will rise to 629 million [2]. In Africa the prevalence of DM will continue to rise, thus imposing an extra burden on a health care system [3]. Mortality and morbidity in patients with DM is result from both the micro and macro-vascular complications. One common complication of diabetes mellitus is erectile dysfunction (ED) [4]. ED is one of the common yet an underestimated complication of diabetes mellitus [5]. Erectile dysfunction is the inability to achieve and maintain an erection sufficient to permit satisfactory sexual intercourse [6]. It may result from psychological, neurologic, hormonal, arterial impairment or from a combination of these factors [7].

The pathophysiology of ED in DM is related to multiple mechanisms including endothelial dysfunction, accumulation of advanced glycation end products, oxidative stress and neuropathy [5]. Diabetes mellitus promotes the onset of ED via vasculopathy from endothelial dysfunction and autonomic neuropathy [8]. In addition, diabetes may affect the cavernous nerve terminals and endothelial cells, resulting in a deficiency of neurotransmitters [9]. It is estimated that the global prevalence of ED should reach 322 million by 2025 [10]. In a study conducted on Ethiopian men with diabetes, 79% of patients never complained about ED [11].

Large differences have been reported on prevalence of erectile dysfunction in different studies. For instance, it has been reported that 49% England [12], 35.8% in Italy [13], 65.4% in Korean [14], 86.1 in Saudi [15], 31% in Kuwait [16], 38.9 % in India [17], 75.0% in Bénin [18], 77.1% south Africa [19], and 67.9% in Ghana [20]. The risk factors for ED are multifactorial and complex. Studies suggest that risk factors for erectile dysfunction in patients with diabetes mellitus included hypertension[16, 18, 19, 21, 22], heart disease [21], cigarette smoking [16, 19, 21-23], low education level[16], increasing body mass index [16, 17, 20], poor glycemic control [16, 18, 20, 24], age above or equal to 50 years [16-18, 20, 23, 25], increasing duration of diabetes [18, 25], presence of depressive symptom [17], high income [20] fat-rich diets[26], and other diabetic complications [27].

Patients with erectile dysfunction might suffer from poor quality of life[22, 28], anxiety when sexual ability declines[29], mutual mistrust, general unhappiness, and fear of losing support from partner [30]. It is known that the good control of the disease is associated with reduced complications[18]. According to literature evidence about one-third of men with erectile dysfunction has improvement in sexual function based on lifestyle interventions, such as diet, exercise and weight loss, cessation of smoking, counseling, and appropriate glycemic control through diet [31, 32]. Different primary studies in Africa showed the magnitude of erectile dysfunction. However, variation among those studies was seen. Therefore, this systematic review and meta-analysis was aimed, to estimate the pooled prevalence of erectile dysfunction patients with diabetes mellitus and its association with body mass index and glycated hemoglobin in Africa.

## METHODS

### Data sources and literature search strategy

Electronic databases such as PubMed, Google Scholar, Africa journal of online, Scopus, Web of science, Psyinfo and Cochrane library were searched by authors independently and systematically. In addition, a manual search of gray literature and other related articles were deployed to identify additional relevant research. Data from International Diabetic Federation (IDF) was searched and used. This search employed articles published from 1^st^ January 1990 to 4^th^ September/ 2019. The search was restricted to the full text, free article, study category of human and English language publications. Likewise, authors were communicated for full text through email. The search was conducted using the following terms and phrases: “erectile dysfunction”, “sexual dysfunction”, “impotence” “diabetes mellitus”, and “Africa”. Boolean operators like “AND” and “OR” were used to combine search terms. Particularly, to fit advanced PubMed database, the following search strategy was used (“erectile dysfunction” OR impotence OR “sexual dysfunction”) AND (“diabetes mellitus”) AND (Africa).

### PECOS guide

#### Type of participants

This review was considered studies that included adult male patients aged 18 or above, and who have been diagnosed with a DM.

#### Type of exposure

Study participants who had BMI≥30kg/m^2^ and glycated hemoglobin greater than 7%

#### Comparison

Study participants BMI 18.5 – 24.9 kg/m^2^ and glycated hemoglobin less than 7%

#### Study Outcome

The outcome of this study was the prevalence of erectile dysfunction among men with diabetes mellitus

#### Types of studies design

The systematic review was included studies done using observational designs such as retrospective or prospective cohort studies, cross-sectional and case control, where erectile dysfunction among diabetic mellitus patients have been reported.

### Eligibility criteria

Studies were included in the meta-analysis if they: (1) All observational studies, which reported the prevalence of erectile dysfunction;(2) articles published in peer reviewed journals and gray literature:(3) published in the English language from 1990 to 2019;and (4) studies conducted in Africa, which included male participants. Studies were excluded if: (1) studies which were not fully accessed; (2) studies with duplicated citation; (3) studies with poor quality score as per stated criteria; (4) articles in which fail to determine the outcomes (erectile dysfunction); and (5) studies included only females.

### Selection and quality assessment

Two independent investigators were screened the title and abstract. Data were extracted using standardized data extraction format prepared in a Microsoft excel by three independent authors. For each article, we extracted data regarding the name of author’s, year of publication, study area, study design, sample size, data collection year, sampling technique, diagnostic criteria used for erectile dysfunction, reported prevalence with its 95% confidence interval and information regarding the associated factors. When some information was missing, first or corresponding authors of the article were communicated at least twice in a month to obtain the variables of interest. The quality of each included study was assessed using Newcastle-Ottawa scale [33]. Representativeness of the sample, response rate, measurement tool used, comparability of the subject, appropriateness of the statistical test used to analyze the data are some of the key criteria in Newcastle–Ottawa scale. Studies were included in the analysis if they scored ≥5 out of 10 points in three domains of ten modified NOS components for cross-sectional study[34]. Any disagreements at the time of data abstraction were resolved by discussion and consensus (Supplementary file 1).

### Statistical analysis

To obtain the pooled effect size, a meta-analysis using random-effects DerSimonian and Laird model was performed [35, 36]. Heterogeneity across the included studies was checked using the chi-square based Q test and the I^2^ statistics test with a value ≥75% for the first and P < 0.05 indicating the presence of significant heterogeneity [36]. To investigate the sources of heterogeneity meta-regression and sub group analyses were performed. Potential publication bias was assessed by visual inspection of a funnel plot. In addition, Egger regression test was conducted and a p□≤ 0.05was considered statistically significant for the presence of publication bias [37]. Sensitivity analysis was deployed to see the effect of single study on the overall effect estimation. The meta-analysis was performed using the STATA version 14 statistical software for windows [38].

### Data synthesis and reporting

To estimate the overall prevalence of erectile dysfunction in patients with diabetes mellitus, the Preferred Reporting Items for Systematic Reviews and Meta-Analyses (PRISMA) guideline was used [39]. The entire process of study screening, selection and inclusion were described with the aid of a flow diagram. Results were presented using forest plots and summary tables (Supplementary file 2-PRISMA checklist).

## Result

### Search results

In the first step of our search, 1,622 studies were retrieved. About 1,617 studies were found from seven international databases and the remaining 5 were through manual search. Databases includes; PubMed (43), Scopus (28), Google scholar (800), web of science (317), Cochrane library (3), Psyinfo (19) and Africa online journal (407). Of these, 879 duplicate records were identified and removed. From the remaining 743 articles, 677 articles were excluded after reading of titles and abstracts based on the pre-defined eligible criteria. Finally, 66 full text articles were assessed for eligibility criteria. Based on the pre-defined criteria and quality assessment, only 20 articles were included for the final analysis (Fig. 1)

**Figure 1.**
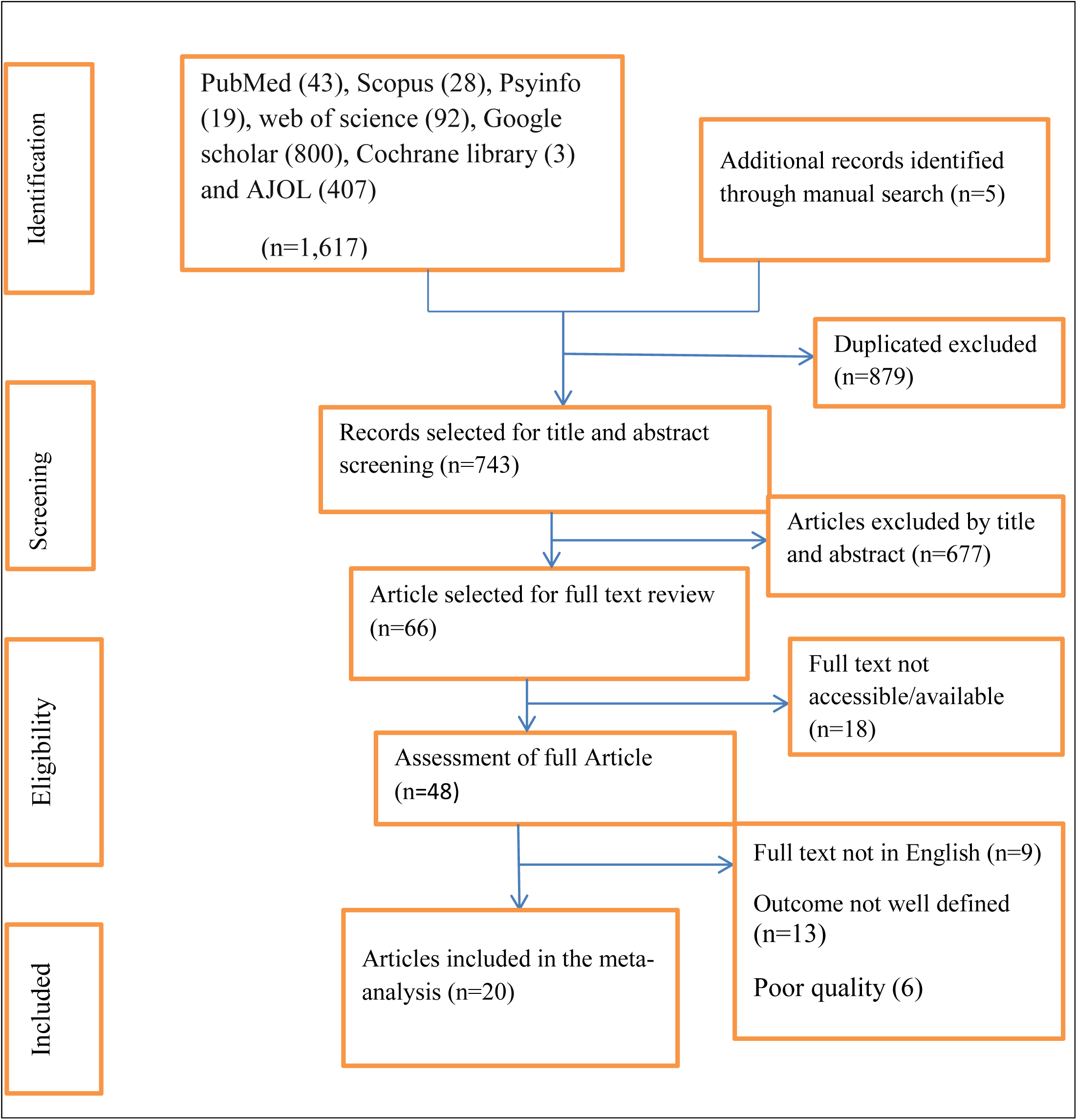
PRISMA flow chart of study selection.

### Baseline characteristic of the included studies

In the current meta-analysis, a total of 20 studies with 5,177 study participants were included to estimate the pooled prevalence of erectile dysfunction among DM patients. Concerning study design, majority (95%) of the studies are cross-sectional. The studies varied substantially in sample size ranged from 70 to 599. The highest prevalence (95%) of erectile dysfunction was reported in a study conducted in South Africa[40], whereas the lowest prevalence (15.5%) was reported in a study conducted in Zimbabwe[41]. Overall information regarding the prevalence was obtained from various countries in Africa. Three of the studies were from Nigeria [42-44], three from Ethiopia[11, 45, 46], two from Zimbabwe[41, 47], two from South Africa [40, 48], one each from Guinea[49], Benin[18], Morocco[50], Egypt[24], Senegal[51], Kenya [52], Ivory coast [53], Tanzania[54], Uganda[55],and Ghana[20].

Regarding the sampling technique employed, in eleven of the studies[11, 20, 24, 41, 43, 44, 47, 50, 52, 53, 55] were used consecutive sampling technique to select study participants. However, two studies [49, 54] did not report the sampling methods used. Concerning tools, ten studies [24, 43-47, 49, 52-54] were used international index of erectile function, two [40, 48] used the sexual health inventory for men, one study [20] used golombok rust inventory of sexual satisfaction and seven studies[11, 18, 41, 42, 50, 51, 55] did not report the tool used to estimate the prevalence of erectile dysfunction patients with DM. The quality score of each primary study, based on Newcastle Ottawa quality score assessment all 20 article have moderate to high quality score (Table 1).

**Table 1.** Shows the baseline characteristic of the included studies.

| First Author | Publication Year | Country | study design | sample size | prevalence (95% CI) | Data collection year | Data Collection Tool | Sampling Technique | Quality score (10) |
| --- | --- | --- | --- | --- | --- | --- | --- | --- | --- |
| Amanyam C etal [42] | 2016 | Nigeria | Cross-sectional | 178 | 45.1(37.8,52.4) | NR | NR | systematic random | **7** |
| Balde N etal [49] | 2006 | Guinea | Cross-sectional | 187 | 48(40.8,55.2) | NR | International Index of Erectile Function | NR | **7** |
| Djrolo, F.etal [18] | 2014 | Benin | cross-sectional | 150 | 75(68,81.9) | NR | NR | Systematic random | **6** |
| El Achhab Y etal [50] | 2008 | Morocco | Cross-sectional | 89 | 82(74,89.9) | NR | NR | Consecutive | **7** |
| El Saghier EO et al [24] | 2015 | Egypt | cross-sectional | 70 | 85.7(77.5,93.9) | March 2014 to January 2015. | International Index of Erectile Function-5 | Consecutive | **7** |
| Gueye S etal [51] | 1998 | Senegal | Cross-sectional | 431 | 16(12.5,19.5) | NR | NR | simple random | **8** |
| Kemp T etal[40] | 2015 | South Africa | cross-sectional | 150 | 95(91.5,98.5) | NR | The Sexual Health Inventory for Men | simple random | **7** |
| Likata GMU etal [52] | 2012 | Kenya | cross-sectional | 350 | 68.8(63.9,73.6) | NR | International Index of Erectile Function | Consecutive | **7** |
| Lokrou A etal [53] | 2011 | Ivory coast | Cross-sectional | 112 | 74.1(65.9,82.2) | NR | International index of erectile function | Consecutive | **7** |
| Mutagaywa RK etal [54] | 2014 | Tanzania | cross-sectional | 312 | 55.1(49.5,60.6) | May to December 2011 | International Index of Erectile Function | NR | **7** |
| Nambuya .A etal [55] | 1996 | Uganda | cross-sectional | 252 | 22.2(17,27.3) | 1 january 1993 to 10 august 1994 | NR | Consecutive | **8** |
| Olarinoye J etal [43] | 2006 | Nigeria | Cross-sectional | 77 | 74(64.2,83.8) | NR | International Index of Erectile Function | Consecutive | **7** |
| Owiredu WK et al [20] | 2011 | Ghana | cross-sectional | 300 | 67.9(62.4,73.3) | November, 2010 to March, 2011. | Golombok Rust Inventory of Sexual Satisfaction | Consecutive | **7** |
| Pasipanodya Ian etal [47] | 2018 | Zimbabwe | cross-sectional | 348 | 73.9(69.3,78.5) | 23 October 2013 to 09 July 2014 | International Index of Erectile Function | Consecutive | **6** |
| Saravoye T [41] | 2016 | Zimbabwe | Cross-sectional | 284 | 15.5(11.3,19.7) | NR | NR | consecutive | **7** |
| Seid A et al.[45] | 2017 | Ethiopia | cross-sectional | 249 | 69.9(64.2,75.6) | January 1 – February 30, 2016 | International Index of Erectile Function | systematic random | **8** |
| Seyoum B [11] | 1998 | Ethiopia | Cross-sectional | 292 | 48.7(42.9,54.4) | NR | NR | Consecutive | **6** |
| Ugwumba FO etal [44] | 2018 | Nigeria | Cross-sectional | 325 | 94.7(92.2,97.1) | September 2016 to December 2017 | International Index of Erectile Function | Consecutive | **7** |
| Walle B | 2018 | Ethiopia | Cross-sectional | 422 | 85.5(82.1,88.8) | January to March, 2016 | international index of erectile function | systematic | **6** |
| Webb EM etal [48] | 2014 | South Africa | cluster randomize control trial | 599 | 36(32.1,39.8) | NR | sexual health inventory for men | Systematic | **8** |

### Prevalence of erectile dysfunction

The finding of the current meta-analysis using random effects model showed that the estimated overall prevalence of erectile dysfunction patients with diabetes mellitus was 61.62% (95% CI: 48.35, 74.90) with high significant level of heterogeneity (I^2^ = 83.4%; p<0.001) (Fig. 2).

**Figure 2.**
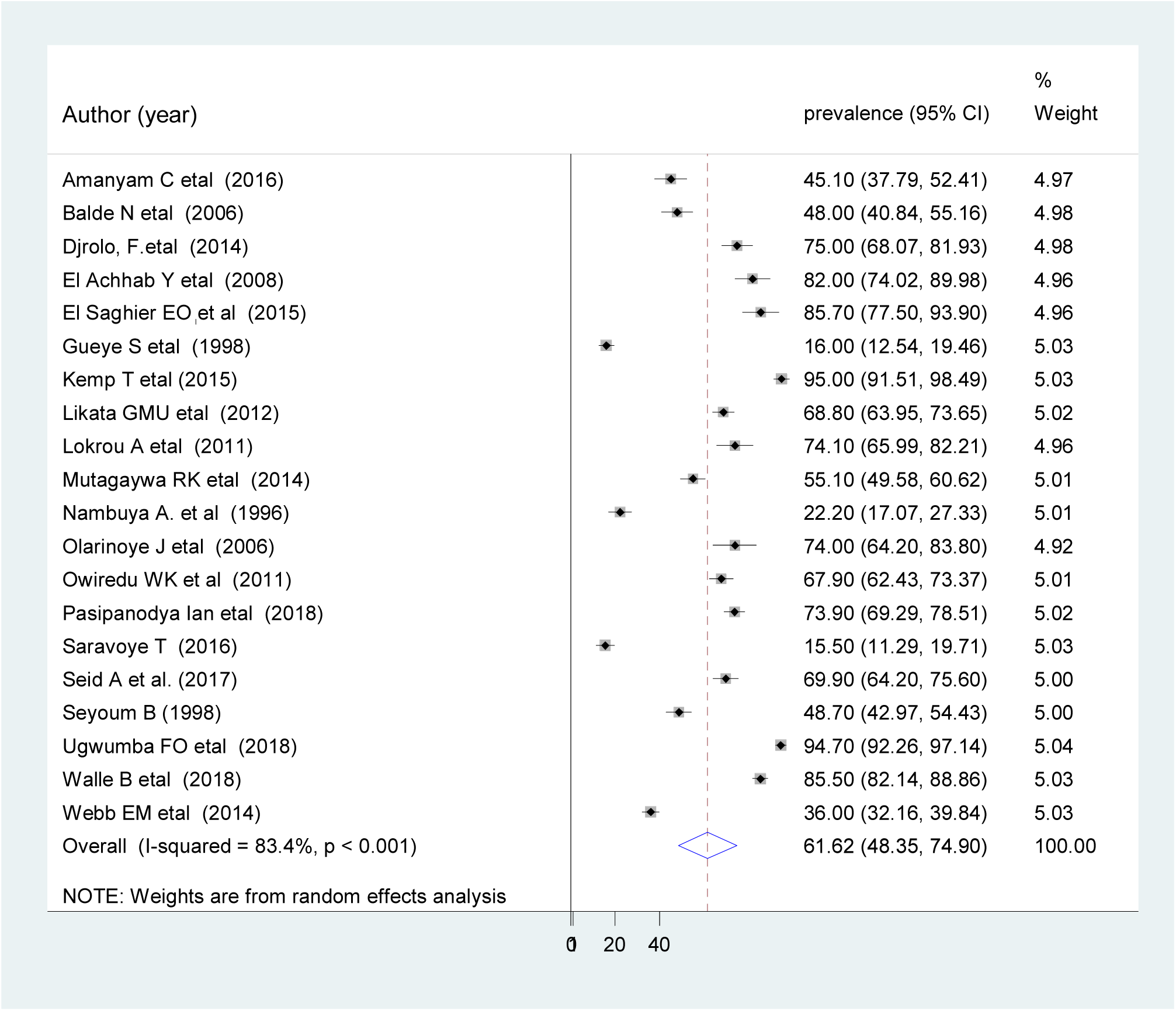
Forest plot of the prevalence of erectile dysfunction.

### Subgroup analysis

To overcome the presence of heterogeneity, subgroup analysis using country, publication year, sample size, and sampling technique was done. Based on this, the prevalence of erectile dysfunction found to be 71.37% in Nigeria, 67.29% in publication year above 2010, and 62.24% sample size greater or equal to three hundred (Table 2).

**Table 2.** The prevalence of erectile dysfunction and heterogeneity estimate with a p-value for the subgroup analysis

| Sub-group |  | No of study included | Sample size | Prevalence (95%CI) | p-value | I<sup>2</sup> |
| --- | --- | --- | --- | --- | --- | --- |
| By year of publication | Before 2010 | 6 | 1,337 | 48.25(27.76, 68.73) | <0.001 | 81.2 |
|  | 2010 and above | 14 | 3,849 | 67.29(53.48, 81.11) | <0.001 | 86.6 |
| Sample size | <300 | 12 | 2,001 | 61.21(43.35, 79.06) | <0.001 | 82.8 |
|  | ≥300 | 8 | 3,087 | 62.24(40.77, 83.70) | <0.001 | 85.3 |
| Country | Nigeria | 3 | 503 | 71.37(38.32,104.42) | <0.001 | 86.8 |
|  | South Africa | 2 | 749 | 65.51(7.9,123.32) | <0.001 | 83.7 |
|  | Ethiopia | 3 | 963 | 68.12(46.28,89.97) | <0.001 | 86.4 |
|  | Zimbabwe | 2 | 632 | 44.69(-12.54,101.12) | <0.001 | 82.7 |
|  | Others | 10 | 2,241 | 59.37(42.19,76.56) | <0.001 | 78.4 |
| Sampling technique | Systematic | 5 | 1,598 | 74.11(57.58, 90.65) | <0.001 | 72.7 |
|  | Simple random | 2 | 501 | 50.74(-17.56,119.05) | <0.001 | 89.3 |
|  | Consecutive | 11 | 2,779 | 59.45(43.54,75.35) | <0.001 | 85.2 |
|  | Not specified | 2 | 299 | 60.97(35.40, 86.55) | <0.001 | 76.1 |

### Meta-regression analysis

To identify source of heterogeneity meta-regression was undertaken by considering year of publication, sample size and sampling technique. However, the result of meta regression showed that both covariates were not statistically significant for the presence of heterogeneity (Table 3).

**Table 3.**
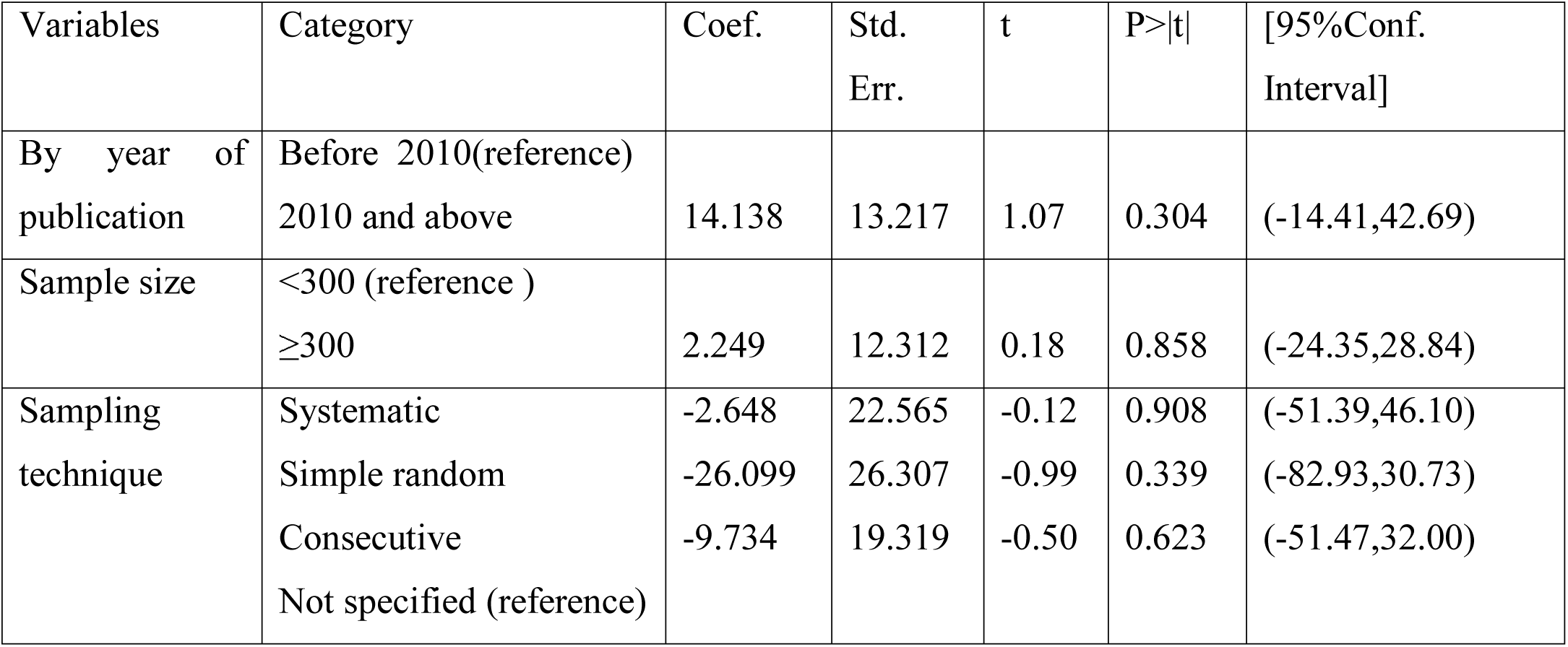
Meta-regression analysis for the included studies to identify source of heterogeneity

### Sensitivity analysis

To evaluate the effect of individual study on the pooled effect size sensitivity analysis was conducted. The finding of sensitivity analyses using random effects model revealed that no single study influences the overall prevalence of erectile dysfunction (fig. 3).

**Figure 3.**
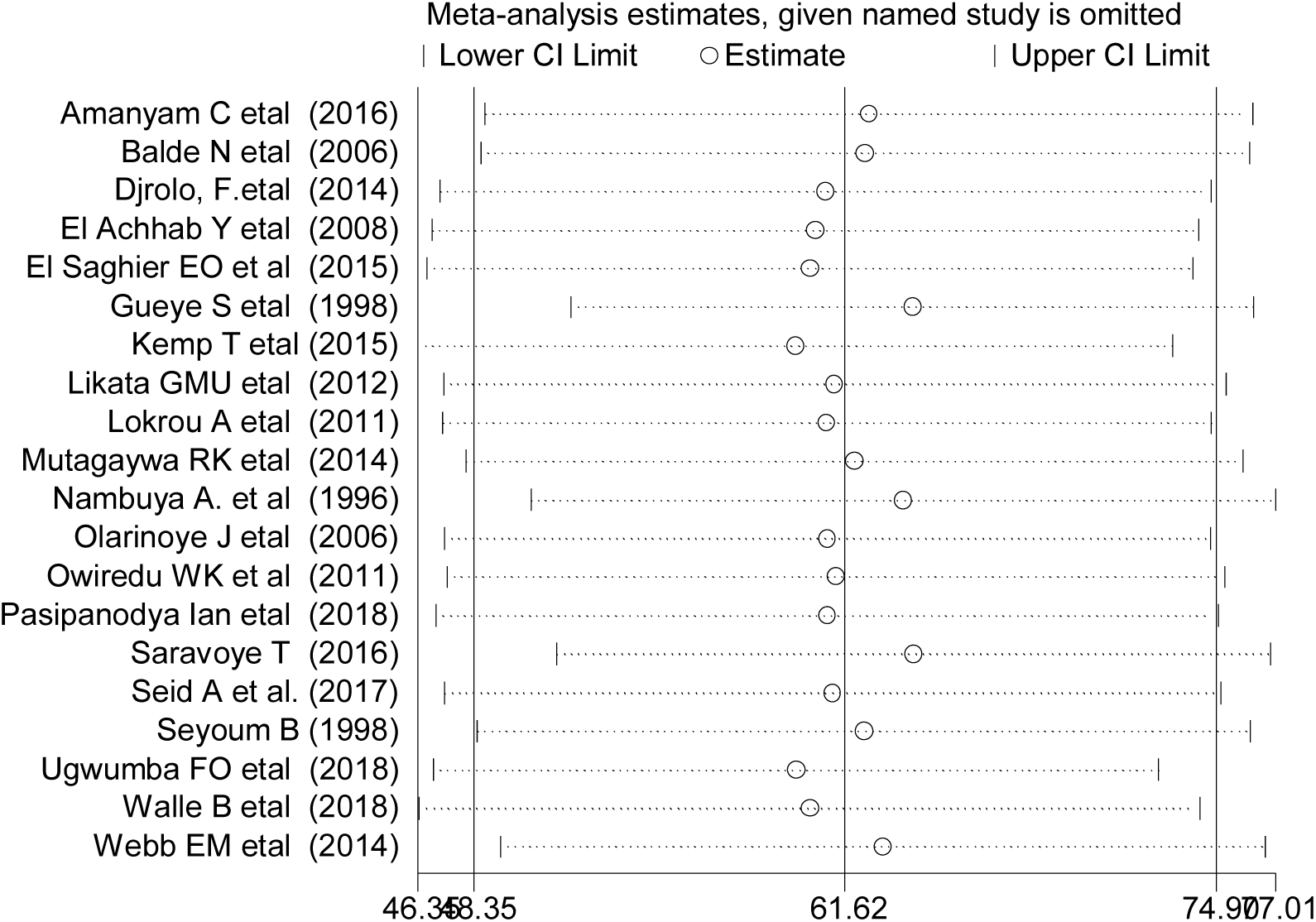
Result of sensitivity analysis of the 20 studies.

#### Publication bias

A funnel plot showed a symmetrical distribution (Fig. 4). Likewise, Egger’s regression test (p=0.406) which indicated the absence of publication bias.

**Figure 4.**
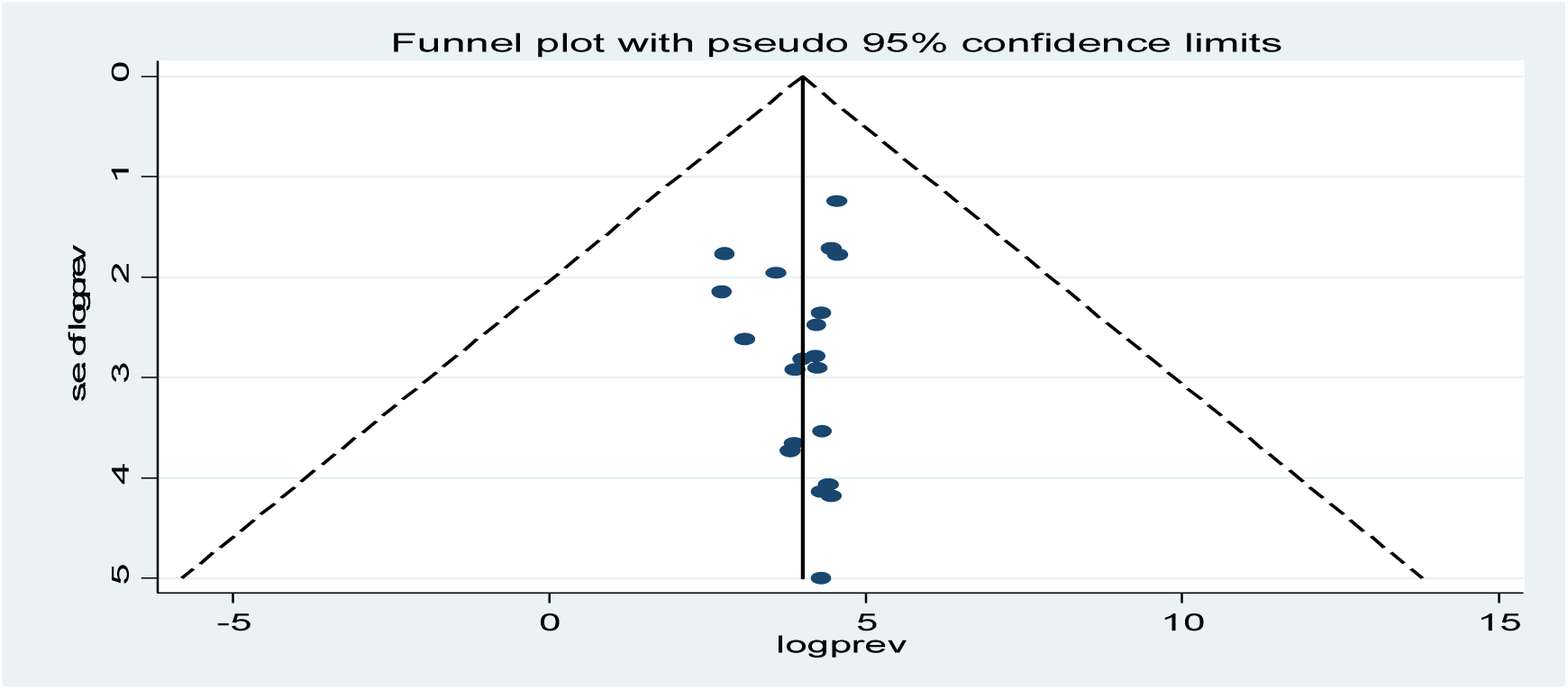
Funnel plot the presence of publication bias among 20 included studies.

#### Body mass index

The pooled effects of five studies showed that body mass index ≥ 30kg/m^2^ was not statistically associated with erectile dysfunction (Fig. 5). The heterogeneity test (I^2^= 89.5%) shows significant evidence of variation across studies. The evidence from Egger’s test shows that there was a no publication bias (P = 0.807).

**Figure 5.**
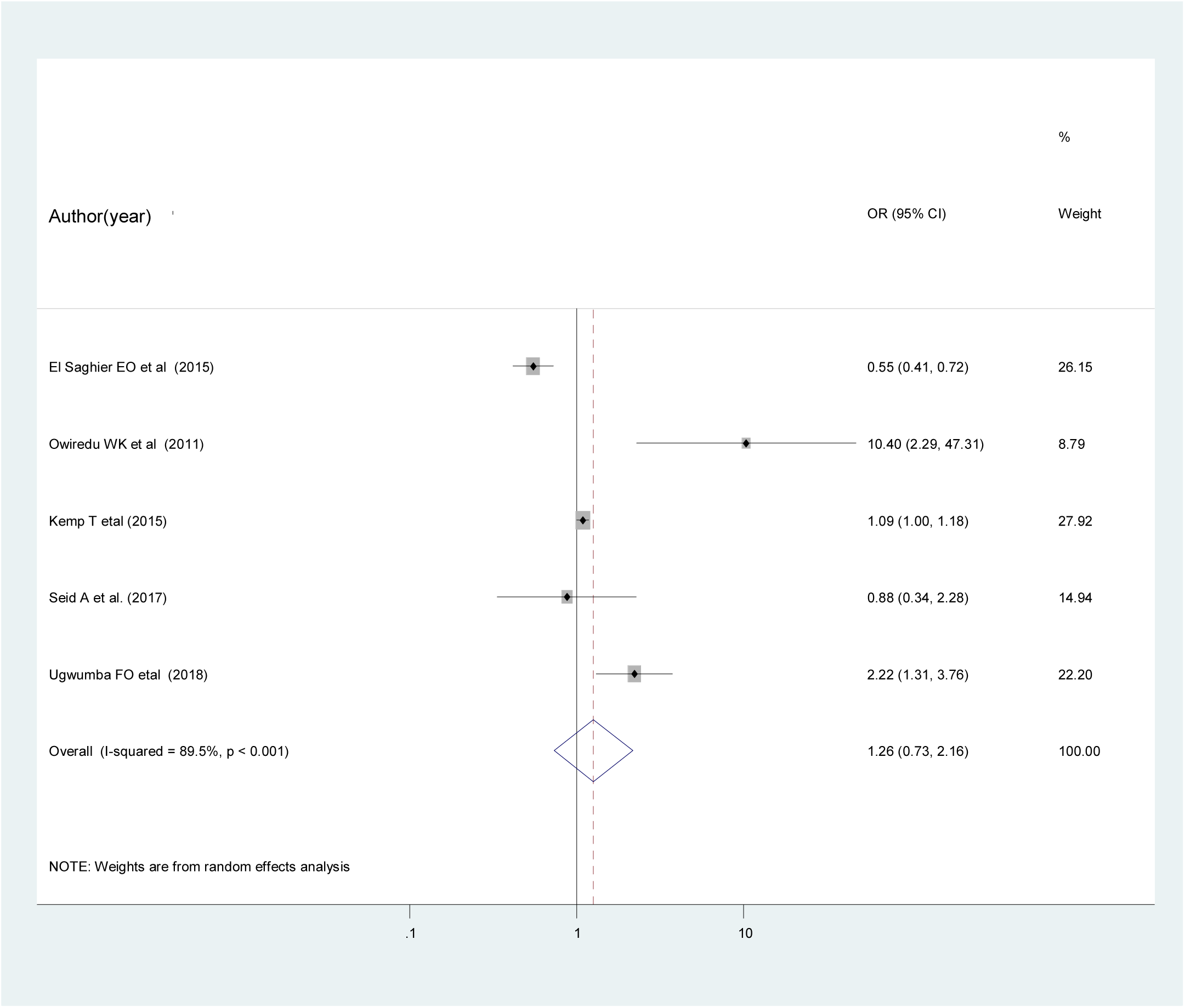
The pooled effects of overweight on erectile dysfunction.

#### Glycosylated hemoglobin (Hga1c)

According to the recent meta-analysis, those who had normal glycated hemoglobin were 7% less likely to develop erectile dysfunction compared to those who had glycated hemoglobin value ≥ 7% although not statistically significant (OR: 0.93 (95% CI (0.15, 5.91)) (Fig. 6). The heterogeneity test (I^2^= 94.4%) showed a significant evidence of variation across studies. The result of Egger’s test showed that no evidence of publication bias (P = 0.147).

**Figure 6.**
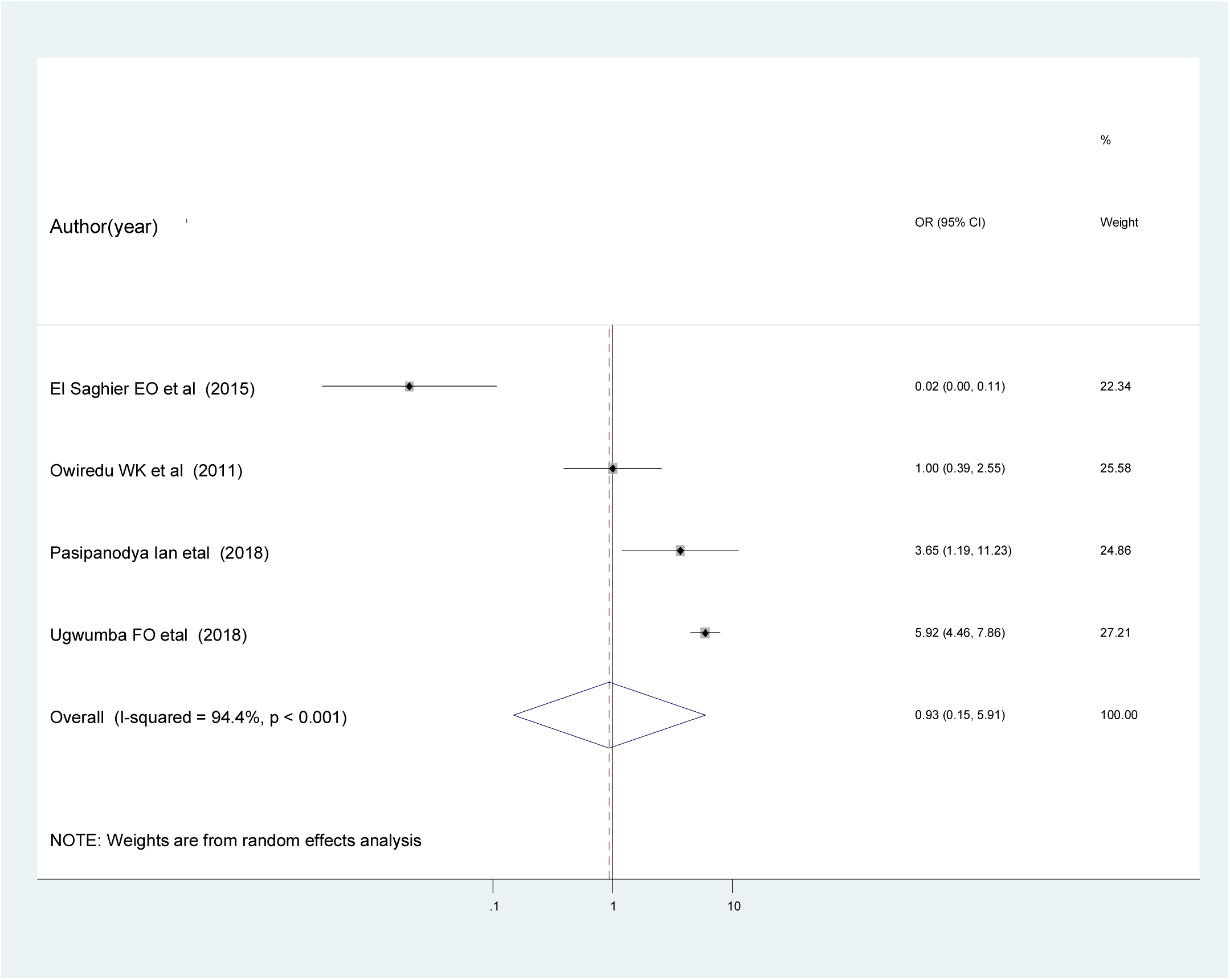
The pooled effects of glycated hemoglobin ≥7% on erectile dysfunction.

## Discussion

The prevalence of erectile dysfunction in this meta-analysis estimated 61.62 % in patients with diabetes mellitus. This implies that there is a high magnitude of erectile dysfunction among diabetic patients, which shows inadequate control of diabetes mellitus related complication. Hence, serious and multifactorial approach is required to manage the diabetic related complications, which emphasis on the treatment adherence, screening for complication, encourage on self-care practice, and health information dissemination.

In comparison with other similar studies, this estimated prevalence of erectile dysfunction in Africa seems significant higher than report from a systematic review and meta-analysis conducted at global level that implies the prevalence of erectile dysfunction among diabetic patients was 52.5% [56]. This variation could be justified by the different diagnostic criteria of erectile dysfunction, the age of the participants in each study, duration and severity of diabetes in their participants.

The subgroup analysis of this study showed that the pooled prevalence of erectile dysfunction among diabetic patients in Nigeria was 71.37% (95% CI: 38.3, 104.4) which is higher than the prevalence in Ethiopia 68.12% (95% CI: 46.3, 89.9), South Africa 65.5% (95% CI; 7.9, 123.3), and Zimbabwe 44.69% (95% CI: 12.5, 101.1). This variation might be due to difference in health seeking behavior between the populations, differences in the diagnostic methods and socio-demographic characteristics.

Though we have identified two factors (body mass index and glycated hemoglobin), to conduct the pooled analysis of two or more AOR of studies, BMI ≥ 30 kg/m^2^ and glycated hemoglobin ≥7% were not associated with erectile dysfunction among diabetic patients. Those who had BMI ≥ 30 kg/m^2^ were 26% more likely to develop erectile dysfunction. Thought it is not statistically significant. This finding supported by other studies conducted in Vietnam [57] and Brazil [58]. Those who had glycosylated hemoglobin < 7% were 7% less likely to develop erectile dysfunction. Thought it is not statistically significant. However, this finding supported by other studies conducted in Korean[14] and china [59].

This study has an implication for clinical practice and patients. Estimating the overall prevalence of erectile dysfunction provide current evidence to design preventive measure, to improve patient quality of life, early detection of poorly controlled diabetic mellitus, close monitoring of poor patient progress and management strategy for ED. In addition, identifying associated factors help health care professional to consider during their clinical care.

Although this meta-analysis conducted with the use of comprehensive search strategy to incorporate the studies there are some limitations that need to be considered in the future research: First, It may be lacked representativeness because no data were found from the entire continent; second, only English articles were considered; and third, this study do not identify all the predictors of erectile dysfunction among patients with diabetes mellitus. Therefore, we would like to recommend the conduct of further study using similar diagnostic tool to identify associated factors for the development of erectile dysfunction among patient with diabetes mellitus.

## Conclusion and recommendations

This study revealed that the prevalence of erectile dysfunction was remains high in Africa. Besides, the prevalence of erectile dysfunction differed by country. Therefore, situation based interventions and country context specific preventive strategies could be developed to reduce the magnitude of erectile dysfunction among patients with diabetes mellitus.

## Data Availability

The data analyzed during the current meta-analysis is available from the corresponding author on reasonable request.

## Abbreviations

AOR: Adjusted odd ratio
CI: Confidence Interval
DM: diabetes mellitus
ED: Erectile Dysfunction
IDF: International Diabetic Federation
PRISMA: Preferred Reporting Items for Systematic Reviews and Meta-Analyses.

## Declaration

### Ethics approval and consent to participate

Not applicable.

### Consent for publication

Not applicable.

### Authors’ contributions

WSS and TYA developed the protocol and involved in the design, selection of study, data extraction, and statistical analysis and developing the initial drafts of the manuscript. YAA and TYA involved in data extraction, quality assessment, statistical analysis and revising. WSS and YAA prepared the final draft of the manuscript. All authors read and approved the final draft of the manuscript.

## Acknowledgements

Not applicable

